# Implementation of a digital early warning score (NEWS2) in a cardiac specialist and general hospital settings in the COVID-19 pandemic: A qualitative study

**DOI:** 10.1101/2022.06.08.22276134

**Authors:** Baneen Alhmoud, Timothy Bonnici, Riyaz Patel, Daniel Melley, Louise Hicks, Amitava Banerjee

## Abstract

**Objectives:** To evaluate implementation of EHR-integrated NEWS2 in a cardiac care setting and a general hospital setting in the COVID-19 pandemic.

**Design:** Thematic analysis of qualitative semi-structured interviews with purposefully sampled nurses and managers, as well as online surveys.

**Settings:** Specialist cardiac hospital (St Bartholomew’s Hospital) and General teaching hospital (University College London Hospital).

**Participants:** Eleven nurses and managers from cardiology, cardiac surgery, oncology, and intensive care wards (St Bartholomew’s) and medical, haematology and intensive care wards (UCLH) were interviewed and sixty-seven were surveyed online.

**Results:** Three main themes emerged: (i) Implementing NEWS2 challenges and supports; (ii) Value of NEWS2 to alarm, escalate, particularly during the pandemic; and (iii) Digitalisation: EHR integration and automation. The value of NEWS2 was partly positive in escalation, yet there were concerns by nurses who undervalued NEWS2 particularly in cardiac care. Challenges, like clinicians’ behaviours, lack of resources and training and the perception of NEWS2 value, limit the success of this implementation. Changes in guidelines in the pandemic have led to overlooking NEWS2. EHR integration and automated monitoring are improvement solutions that are not fully employed yet.

**Conclusion:** Whether in specialist or general medical settings, the health professionals implementing EWS in healthcare face cultural and systems related challenges to adopting NEWS2 and digital solutions. The validity of NEWS2 in specialised settings and complex conditions is not yet apparent and requires comprehensive validation. EHRs integration and automation are powerful tools to facilitate NEWS2 if its principles are reviewed and rectified, and resources and training are accessible. Further examination of implementation from the cultural and automation domains are needed.

## Introduction

Prediction tools in acute care settings can improve patient safety through enhanced efficiency of care and reduced pressure on health systems. Early Warning Scores (EWS) are a potential solution to decrease critical events, unnecessary deaths and debilitating resources. EWS have become part of the escalation guidelines directing clinicians to the level of care needed. In conjunction, clinicians utilise their education and clinical experience, as when EWS did not exist, to make clinical judgments. Implementing EWS advise clinical assessment when puzzles are missing from knowledge and experience.

However, there is a gap in evidence on the performance of EWS in different settings and specialities. (1–4). For instance, in cardiac care and complex comorbidities, i.e. COVID-19 patients, the performance of EWS are poorly and in the early stages. Equally, there is a lack of evidence on implementing integrated EWS in Electronic health records (EHR) in specialised clinical settings. With the electronic assessment recording, EWS scores and alarms are produced automatically, facilitating its utilisation by clinicians. The functionality gives more confidence in EWS generated when part of the burden becomes the role of the machine. It has improved clinical outcomes and staff workflow (5)

For EWS to be successful, they have to be executed accurately. Errors in assessment, recording and escalation of care contributed to 20-80% of the severe adverse events (6). As shown in previous EWS and digital solutions, wide dissemination does not necessarily lead to successful adoption. (7). It is well established that failure of EWS are related to patients’ physiology or professionals’ practice, i.e. poor adherence to the prescribed protocol for deterioration (7,8). In addition, the downsides of automated monitoring, i.e. measurements errors, artefacts and false alarms(9) can challenge the progress needed. The significance of NEWS2 and automated application is only valuable if resulting in a tangible improvement.

Therefore, the implementation of NEWS2 requires investigation by understanding clinicians’ perceptions of the application. We conducted a qualitative study of EHR-integrated NEWS2 in a specialised cardiac; and a general hospital; from the perception of nurses utilising it. The non-adoption, abandonment, scale-up, spread, sustainability (NASSS) tool (10) is a pragmatic, evidence-based design that can provide a thorough understanding of digitally supported tools in healthcare. Due to the application of electronic recording and automation, we followed the NASSS framework in the study.

### Previous implementation studies

EWS models that proceeded NEWS2 were examined from nurses’ and doctors’ experiences in acute and non-acute settings. In a study in Norway, MEWS2 supported early recognition and knowledge sharing between nurses (11). Another study found that nurses value NEWS as it incorporates their knowledge and judgment, yet may not necessarily lead to desired clinical outcomes (12). In non-acute settings, it is believed to facilitate communication and decision making. However, EWS used in emergency departments (HEWS) was unvalued by physicians and nurses (13). HEWS may not be as advanced as NEWS and NEWS2 in development. Nonetheless, results from specialised departments like ED demonstrates the need to examine settings with unique functionality. The negative experience of NEWS caused tension when it was implemented(14). Compliance, workload pressure and discrepancies between clinical judgment and the scores generate workplace anxiety(7,14). As pressure increases in busy hours, defective collaboration and miscommunication arise between clinicians leading to failed implementation(7). The experience of EHR-integrated EWS in specialised settings is missing from the literature.

### Objectives

To qualitatively evaluate the success and role of implementing EHR-integrated NEWS2 from nurses’ perception in a cardiac specialist and general hospital settings in the COVID-19 pandemic.

## Methodology

### Study settings

We selected two different sites: St Bartholomew’s is a cardiac specialist and teaching hospital in London, and has heart and cancer centres, with other related specialities. University College London Hospital is a general teaching hospital. NEWS was first implemented in 2012, followed by the updated version NEWS2 in 2018 in both hospitals. NEWS shifted from paper version into EHR-embedded format calculated from each vital signs’ measurements, via Cerner™ in Barts and EPIC™ in UCLH. NEWS2 update was reflected in EHRs systems.

➣ NEWS2 pathway in Barts

NEWS2 begins with assessment and vital signs measurement by nurses and nurse assistants via automated monitoring devices (welch Alyn connex spot monitors). Monitors are connected to Cerner™, transmitting measurements directly to patients’ charts. NEWS2 is calculated automatically in the electronic chart; a score is given, then an alarm is shown when a score indicates the need for intervention. Clinicians need to log in to view the score of the patient.

➣ NEWS2 pathway in UCLH

Nurses and nurse assistants do routine vitals measurements. They input their recordings physically into the patient’s chart in EPIC™. The score is calculated automatically, and an alarm will show when the status indicates attention. Nurses and physicians view the score when logged in to their patients’ charts.

### Study framework

We conducted a qualitative study design to evaluate the implementation, following the NASSS framework (10). The NASSS design was chosen for its compatibility with evaluating the adoption of digitalised health systems in health care settings. The factors in the framework were re-phrased to present the process of implementing NEWS2 and to guide structuring questions around the investigated areas. (Figure1)

**Figure 1:**
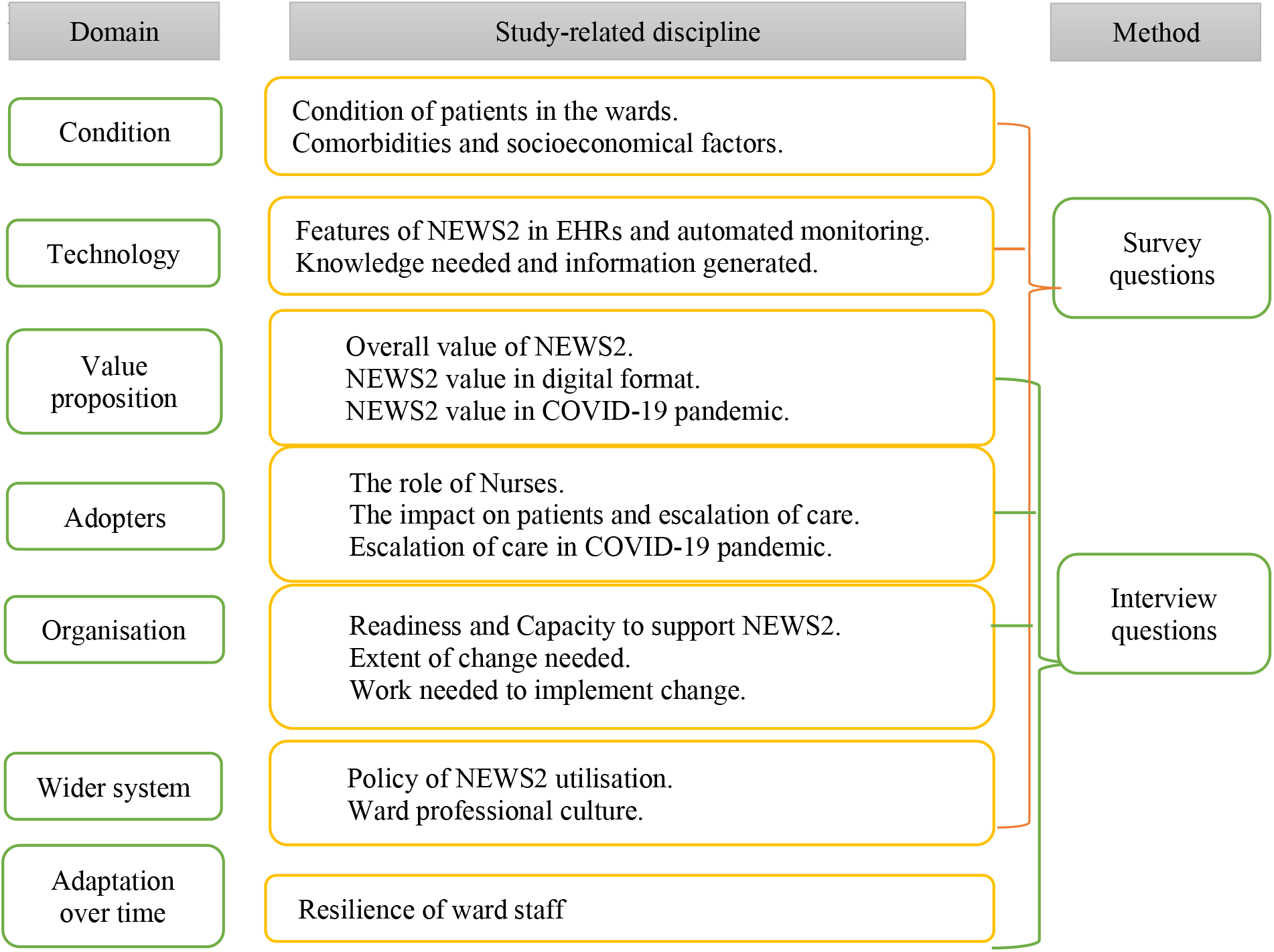
NASS framework domains and methods in the study.

### Data collection

A purposive sampling method was followed with input from Research Team, Critical Care Outreach (CCOT) and Resuscitation Teams in Barts, and Patient Emergency Response Team (PERT) in UCLH, to identify representative participants to contact based on roles and experience in utilising NEWS2. Focus groups were initially planned to gather a collaborative perception of nurses from different hierarchical and role levels: ward nurses and managers.

Invitation emails for focus groups were sent In March 2021 to ward managers and nurses in the cardiac specialist hospital, and a follow-up email was sent ten days later. Due to the workload pressure during the pandemic, assigning participants to the focus group at one time was impractical. Therefore, as discussed with the research team, it was decided to conduct individual interviews (Appendix 2). Invitation emails to online interviews were sent in April 2021 to 10 nurses and managers in Barts, and equally to UCLH staff in June, followed by a reminder after ten days. Information sheets and consents were sent before setting a date for interviews. Informed consent was obtained prior to conducting the interviews.

A questionnaire was created in Smart Survey(15), including consent to answer the survey. A link was sent to nurses and managers in cardiac and non-cardiac wards: Cardiology, Cardiac Surgery, Haematology, oncology wards, and ICU in Barts and Medical, Oncology and Haematology wards, and ICU in UCLH. Wards in UCLH were chosen to provide a mutual environment of patients’ speciality to Barts. Survey questions were matched, excluding the automated monitoring part in UCLH survey (Appendix 1). A reminder was sent after 14 days to boost participation.

### Data analysis

Interviews were recorded in Teams then saved with surveys in the NHS network. Recordings and surveys were pseudorandomised then transferred to UCL Data Safe haven (DSH), a secured database system with restricted access to the PI and CI; via safe gateway technology. Transcription of audio recordings and analysis of transcriptions and surveys were done in Nvivo.

The interviews were analysed thematically to enable us to identify shared ideas and experiences and recognise patterns in datasets(16) following four steps. First, familiarity with the interview by listening to the audio and reading the transcription. Script and audio were compared to achieve reliability. Second, initial codes were assigned to parts of the text with relevancy to research questions. Third, identifying relevant themes and subthemes that captures the idea of significance. Fourth, themes and sub-themes were checked by BA and AB to assess their quality. Fifth, themes were organised and named according to the relativity with the research aim and interest. Discussion with the research team was carried out until agreement was reached on the main themes produced. Results were reviewed and double-checked independently by BA and BA. Finally, the results report comprises four main themes, then exported from DSH.

### Patient and public involvement

Patients and/or the public were not involved in the design, or conduct, or reporting, or dissemination plans of this research.

## Results

11 nurses and managers participated in interviews that lasted 35 minutes each. Survey respondents were 67 nurses and managers.

### Interviews

In the cardiac setting, 6 staff responded, and 4 agreed and were interviewed. In the general hospital, 7 nurses participated: 3 interviewed from the first invite. After follow-up emails, 5 responded, and 4 were interviewed. (Table 1)

**Table 1:** Characteristics of interviews and survey respondents.

|  |  | **Cardiac specialist hospital** |  | **General hospital** |  |
| --- | --- | --- | --- | --- | --- |
|  |  | Interviews | Surveys | Interviews | Surveys |
| *Role* | Manager | 2 | 5 | 4 | 10 |
|  | Nurse | 2 | 20 | 3 | 23 |
|  | Nurse assistant |  | 3 |  | 6 |
| *Speciality* | Cardiology | 4 | 12 |  |  |
|  | Critical care |  | 3 | 2 | 26 |
|  | Medical |  | 4 | 3 | 5 |
|  | ER |  |  | 1 |  |
|  | Oncology/Haematology |  | 11 | 1 | 8 |

### Questionnaires

28 staff answered the surveys in the cardiac setting, from cardiology, critical care, medical and oncology. From the general hospital, 39 answered the questions from critical care, medical and oncology wards. (Table 1)

### Themes

Three themes emerged from applying the NASSS framework on studying the success of implementing NEWS2 in the two hospitals. The themes from domains were as follows: (i)Organisation, wider system, and adaptation over time: Implementing NEWS2 between challenges and supports, (ii)value proposition and adopters: the perceived value of NEWS2 as an alarming tool; in escalation and during the pandemic; (iii) the condition and technology features: EHRs integration and automation of monitoring. Some domains from the framework intersect in themes due to the relativity in subthemes to more than one domain (Figure 2). Results from the survey served as a supplement that supported the themes. Table 2 explains the characteristics of the interviewees hospital setting and digital system.

**Table 2:** Culture and system characteristics of interview participants.

| Participant | Hospital specialization | EHR integration | Automated monitoring | Role | Speciality |
| --- | --- | --- | --- | --- | --- |
| 1S | Cardiac Specialist | √ | √ | Manager | Cardiology |
| 2S | Cardiac Specialist | √ | √ | Manager | Cardiology |
| 3G | General | √ | X | Staff nurse | Cardiology |
| 4G | General | √ | X | Manager | Medical |
| 5G | General | √ | X | Manager | ICU |
| 6G | General | √ | X | Staff nurse | Oncology |
| 7G | General | √ | X | Manager | ICU |
| 8S | Cardiac Specialist | √ | √ | Staff nurse | Cardiology |
| 9G | General | √ | X | Manager | Medical |
| 10G | General | √ | X | Manager | Medical |
| 11G | General | √ | X | Staff nurse | Emergency |

**Figure 2:**
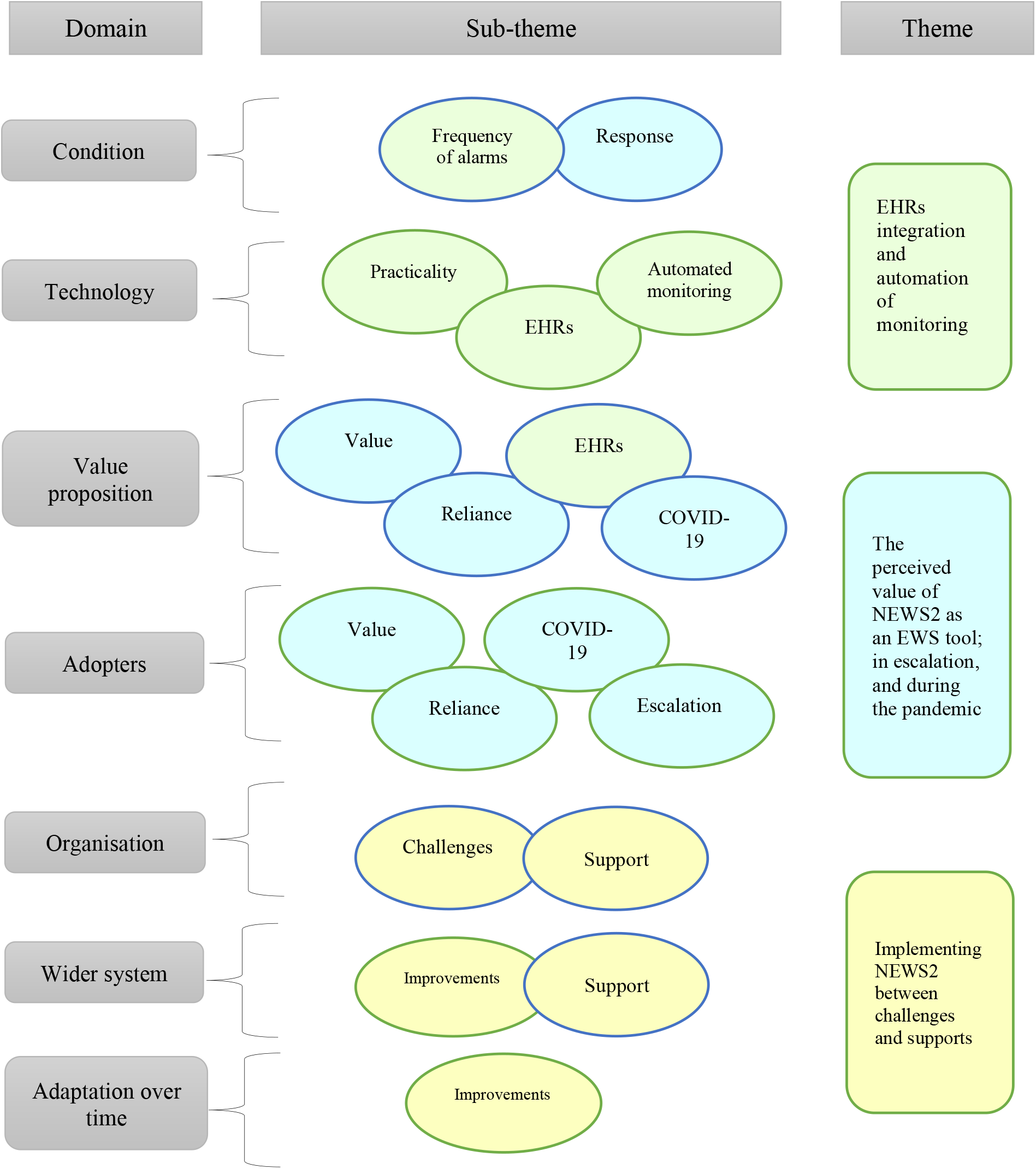
Themes and subthemes emerging from the NASSS framework domains. Note: bubbles colours relate subthemes to the main themes formed.

➣ Implementing NEWS2 between challenges and supports.

The difficulties found by nurses and managers were human-related, tool-related, and resource-related factors.

From the human-centric side, Junior nurses worry if their judgement for escalation based on NEWS2, and their knowledge is inaccurate or undervalued. On the other hand, clinicians and rapid response teams may not always be cooperative when escalation is reported, resulting in timidness by junior staff and avoiding being involved in escalation.

- *I think NEWS2 can be unhelpful when I see how the medical team behave in the situation*.*” 2S*
  - *“It’s about increasing the psychological safe space to speak up no matter who you are. We’re not quite there yet*.*” 7G*

The IT literacy and interest difference between staff cause a gap in adopting NEWS2 in EHRs in both hospitals. Resistance or delays in learning lead to errors in documentation and obstruct escalation.

- “*Documentation isn’t Great*.*” 1S*
- “*Because it’s an electronic system, some staff aren’t particularly comfortable using IT equipment. So, they do the Observation, write on a piece of paper, then enter it later*.*” 9G*

NEWS2 parameters were considered problematic. Their format may not be appropriate for the patient group, specifically cardiac patients, as reported from both sites. There’s a frequent need for parameters adjustment to suit a patient’s medical history to avoid repetitive alarms. Adjustment is challenging as this role is assigned to doctors only.

- “*The problem with it is that the medical team needs to input the target parameters, and only when they do that it does trigger NEWS2*.*” 6G*
- “*Particularly I think cardiology patients need parameter changing*.*” 2S*

Poor resourcing of equipment and staff affects the adoption negatively. When workstations are occupied, recording may be incorrect, and escalation is delayed, yet the issue may not be present in the specialist hospital, where automated monitoring occurs. In addition, nursing assistants who do routine monitoring are not as trained as registered nurses. Reporting deterioration can be missed or delayed.

- “*The healthcare assistants do the observation, then by the time they report, or maybe they forget to tell you the patient is scoring five or six*.*” 8S*
- “*We sometimes don’t have access to an EHR machine because they’re busy or broken, or we haven’t got enough, or we don’t have access to the handheld devices*.*” 10G*

From the support side, nurses and managers had a consensus on the benefits training provided in both sites. However, training has declined due to the pandemic pressure in the workplace. They reported significant support by the hospital management to utilise NEWS2 and adopt the EHR-integrated version, including induction programs training. Ongoing guidance by informatics experts, superusers and ward managers was reported in surveys and interviews. Nonetheless, lack of training and auditing is an issue in both sites. An emphasis was reported on structuring a clear step-by-step process for implementation. Quality projects that focus on improving documentation and escalation, such as deteriorating patient’s dashboard project in the specialist hospital, were valued by staff in both sites.

- “*I think they are supporting it. But I do think it’s a shame that, throughout the pandemic, it’s not audited anymore*.*” 2S*
- *“Other hospitals created dashboards of patients scoring high for users and outreach team to focus on*.*” 7G*

Some nurses in general hospitals perceive it as a mandatory tool rather than a choice yet agree to follow. Resilience was subject to personal and experience differences, such as age and recurrent guidelines updates. Younger staff were reported to be more receptive to change than older staff.

- “*They are quite resilient because there has been much structural process, which has changed constantly, the team have taken them forward quite well*.*” 11G*
- “*They easily pick up on the new electronic things; it’s quite a young team*.*” 6G*
- “*Some don’t change because they find the computer stuff and everything a little bit difficult*.*” 8S*
➣ Value of NEWS2 to alarm, escalate, and in the pandemic.

Overall, nurses and managers believed NEWS2 helps prioritise patients’ needs according to acuity, therefore improving patient safety. They valued the unified language between clinicians to overcome disagreements arising. Tangible advantages of NEWS2 were seen in recognising a response to treatment, a need for transfer to Ward or ICU, or just an impression of the patient’s status. Nonetheless, it is deemed “overvalued” by some nurses, and others anticipated the need for iteration due to its failure in some settings like cardiology and general surgery. Senior staff perceive its usefulness for junior doctors and nurses yet believe it poses the risk of over-reliance in utilising a tool that may not be reliable for each condition. It can restrict their critical thinking due to their lack of experience. They perceive it as an optional mean in the escalation process yet not dependable.

- “*It allows us to catch things before we have to send a patient elsewhere*.*” 1S*
- “*Historically, sometimes nurses will argue if the patient is sicker or not; NEWS2 frames this with a nationally recognised number*.*” 10G*
- “*I used to see patients that were unwell, that didn’t trigger NEWS. Junior nurses worry about what the audit says, they can be completely fixated on a NEWS2 and not the patient holistically. Over-reliance becomes a danger*.*” 7G*

In escalation of care, it provided clarity. When, where and whom to escalate to is coherent to everyone, potentially saving time and promoting safety. Yet, nurses from both sites reported that the impact of NEWS2 in the escalation is insignificant to make a noticeable difference.

- “*This is very clear cut in terms of NEWS2*.*”* 10G
- “*I don’t think it’s made that much of a difference*.*” 11G*

In the time of the pandemic, there was an agreement by most staff that no advantage observed is credited to implementing NEWS2 in clinical work or patients’ outcomes. Nurses and doctors are more vigilant to deterioration due to international and organisational recommendations to manage and prevent COVID-19. Teams, i.e. medical, infection control and CCOT, were present, facilitating the escalation of care. An advantage reported was specific attention to temperature scores in NEWS2 to alert any suspected COVID-19 case.

- “*I don’t think it had much of a value in the pandemic. We had a medical team on our Ward all the time, which we weren’t used to, and of course, we had a good response*.*” 2S*
➣ Digitalisation: EHRs integration and automation.

NEWS2 in EHRs is perceived to have several advantages, with some unpleasing downsides. Accuracy of calculation and timely scoring were reported once entered in patients’ charts. This facilitates decisions for treatment or escalation; and the ability to audit documentation. On the other hand, it has been perceived to inhibit junior nurses’ and doctors’ thought processes when they rely on the system to produce a score without examining its parameters. Some nurses expressed a preference for the paper chart version of NEWS2 over the electronic one due to the absence of colour coding, inability to adjust thresholds, omission of score trend, and constant alarms pop-ups. More dissatisfaction with the model in EHRs was expressed by general hospital staff than Specialist hospital, who agreed more to it. Personal differences like age and IT literacy causes a restrain to some nurses to adopt digital documentation.

- “*We can deep dive in the documentation to make sure that everyone is doing what they need to do*.*” 1S*
- “There are *benefits of an electronic system, but it doesn’t allow nurses and doctors at a junior level to think about their thought processes*.*” 7G*
- “*I did use to like the graphs that we used to get in the paper version, to be able to see what the acuity trend is like for patients and across the floor*.*” 6G*

With automated monitoring in the specialist hospital, the accuracy of recording and timely data transfer is reliable. Nurses are more aware of the need to accomplish this task when it’s automated; not appearing on the screen means undone, while previously it could indicate late entry. Nonetheless, timely observations may not lead to timely escalation. Nurses do not carry computers or handheld devices all the time. Therefore, escalating a case is subjected to completing the documentation on the workstation, which may be by the end of assessing a number of patients.

- “*it’s accurate and timely; the moment you open the screen, it will flash to remind you to act*.*”* 7G
- *“They’ll do a whole lot of patients, six to ten and then come back and escalate it at the end*.*”* 2S

Understanding the information behind NEWS2 and generated by it was straightforward to most participants. Nurses who considered it unideal expressed the need to learn the rationale behind each parameter scoring and confusion related to triggering at the patients’ baseline. Nonetheless, the NEWS2 score, and parameters value is perceived to be unrealistic. Constant unnecessary alarms are disadvantages reported by nurses working in cardiology and oncology wards in both sites, while not alarming when assessment indicates the need to escalate. Nurses who are the primary assessors for NEWS2 are not authorised to adjust the scale, causing annoyance and avoidable alarms when done by doctors.

- *“I suppose cardiology patients need parameter changing. It would help. I think here’s a lot of unnecessary pop-up boxes*.*” 2S*
- *“Sometimes, the patient might not be newsing. But you just know and feel something changed. And often, we’re quite right with that*.*” 7G*.

## Discussion

Our study in the general and specialist settings examined the facilitators, challenges, and value of implementing EHR-integrated EWS guided by the NASSS framework. We have explored nurses’ views in a unique time of the pandemic. The framework’s domains have intersected in the themes leading to three findings identified. The implementation support by hospital decision-makers was sufficient, yet, determined challenges, like clinicians’ behaviours, IT literacy, lack of resources and training and the perception of NEWS2 value, can forbid the success of this implementation. Secondly, the impact of the pandemic on clinical practice and training has resulted in uncontrollable changes and enforcing guidelines that lead to undervaluing NEWS2. Lastly, EHR integration and automated monitoring are strong mediums for improvement that are not fully or precisely employed yet. There was an agreement from both sites on the facilitators and barriers, with preference from the specialist setting of the EHR integration and employing dashboards to improve escalation. The challenges found were cultural and setting related or digital system related, as manifested by participants’ views from both hospitals.

The challenge of seniority-related behaviour can be daunting to junior nurses and doctors that suppress their development in the work setting. Junior clinicians are the most receptive to change in the health system, and their ability to learn is high, a wasted advantage if discouraged. In previous EWS implementation, the seniority level of qualified nurses can affect the response of medical staff to review a patient or not (17). Increasing the safe space to express clinical concerns by junior staff is a major need to be addressed to improve the work culture in hospital settings.

The perception of NEWS2 as a unified EWS for patients with complex conditions is appreciated for patients’ safety and eliminating clinical judgment disagreement, yet insufficiently valued. They doubt their decision to escalate or have a dismissive culture to high NEWS2 score, majorly in the cardiology settings, owing to their clinical knowledge of patients’ baseline and history. Applying an EWS system for critically ill patients can either be a confidence booster if perceived as reliable (11), or a source of tension between their own trusted knowledge and experience and nationally enforced guidelines (18). To date, there is insufficient evidence on the validity of NEWS2 in specialised subgroups, including cardiology and oncology settings (1); therefore, the call for unified NEWS2 remains weak. However, clinicians’ belief toward applying NEWS2 to avoid further risk to patients’ safety is valid.

Inadequate resources and training are challenges to implementing EWS that were heightened during the pandemic. Various medical devices are in shortage globally (19,20) and missed training opportunities created a gap in professional development, negatively impacting clinicians’ confidence.(21,22). The surge in hospitalisation and escalation to ICU due to the COVID-19 pandemic necessitate the enforcement of national and international frameworks as a well emergency response to overcome the crisis (20,23,24). That came ahead of implementing a national EWS developed for ward patients when hospitals were more stable functionally. Greater attention was paid to all patients during the pandemic with the presence of various medical teams, facilitating critical care regardless of NEWS2 score.

The documentation nonadherence presented a cultural issue in both hospitals, with more non-compliance and the need for auditing expressed in the specialist setting. Embedding NEWS2 in EHRs and automated monitoring can be robust solutions representing the role of digitalisation in improving documentation, clinical tasks, and patient outcomes. There is inadequate evidence on the benefit of EHRs integration in previous studies. Our study indicates the advantage of accuracy and timeliness of scoring and alarming NEWS2, prompting decision making and early intervention, and potentially decreasing workload. Automated monitoring has motivated staff to complete documentation since what is seen is done (9). However, digital systems challenges, including insufficient workstations, IT assessment and training(25,26) and overlooking the positive aspects of paper workflow cause transformation to be hindered. Therefore, it is essential to address the obstacles to implementing EWS in EHRs that mimic the challenges of implementing EMRs or digital systems in health settings.

## Strengths and limitations

- The study examined the implementation in two different sites in structure, policies, speciality, and care pathways.
- The NASSS framework was used as a guide, a solid theoretical foundation that analyses the complexity of implementing health technology solutions.
- We conducted the interviews and surveys at the time of the pandemic in England, which provided enhanced rapport and a rich narrative.
- The sample is limited by purposive sampling and the pandemic pressure, which might have restricted further participation in the study.
- The interviews were guided by the domains and may have missed some richness of human-centric topics that could be explored deeply. i.e. seniority behaviours and EHRs users’ preferences.

## Conclusion

The significance of NEWS2 can be underestimated when challenges are overlooked, and evidence of its validation is not apparent. NEWS2 was appreciated partially by some nurses and managers; however, it was not sufficiently strong in specialised care like cardiology to empower the adoption. Clinicians’ behaviour in escalation from a cultural perspective, IT literacy and resources from digitalisation perspective; impact the implementation. COVID-19 related Regulations and guidelines influence clinicians’ practice more than implementing EWS and digital solutions. Implementing new EWS and digital solutions may be less complicated prior to the pandemic. However, more evidence is needed. Studying the validity of NEWS2 in specialised settings and complex conditions is required to guide the implementation. EHRs integration and automation are dynamic tools to facilitate NEWS2 utilisation if the principles of NEWS2 are reviewed and rectified, and resources and training are accessible. There is a need to explore the implementation further from human-centric, cultural, and digital transformation domains.

## Supporting information

SRQR checklist

## Statement

### Contributors

BA, AB and TB conceived the study, BA carried the interviews and surveys with guidance from TB, DM, and LH. Data analysis was conducted by BA with guidance from AB. BA wrote the initial draft of the manuscript and all authors contributed to the interpretation of findings and revision to the manuscript.

### Ethics

This study is registered and approved by the Health Research Authority (HRA) and Health and Care Research Wales (HCRW) and sponsored by University College London (UCL). REC reference: 20/PR/0286.

### Data availability statement

All data relevant to the study are included in the article or uploaded as supplementary information.

### Twitter

@BaneenAlhmoud, @amibanerjee1, @TimBonnici, @DrRiyazPatel, @louisechicks1, @DanBartsICU.

## References

1. Alhmoud B, Bonnici T, Patel R, Melley D, Williams B, Banerjee A. Performance of universal early warning scores in different patient subgroups and clinical settings: a systematic review. BMJ Open. 2021 Apr 1;11(4):e045849.

2. Grant S. Limitations of track and trigger systems and the National Early Warning Score. Part 1: areas of contention. Br J Nurs. 2018 Jun 12;27(11):624–31.

3. Grant S, Crimmons K. Limitations of track and trigger systems and the National Early Warning Score. Part 2: sensitivity versus specificity. Br J Nurs. 2018 Jun 28;27(12):705–10.

4. Russell S, Stocker R, Barker RO, Liddle J, Adamson J, Hanratty B. Implementation of the National Early Warning Score in UK care homes: a qualitative evaluation. Br J Gen Pract. 2020 Nov 1;70(700):e793 LP–e800.

5. Subbe CP, Duller B, Bellomo R. Effect of an automated notification system for deteriorating ward patients on clinical outcomes. Crit Care. 2017;21(1):52.

6. Difonzo M. Performance of the Afferent Limb of Rapid Response Systems in Managing Deteriorating Patients: A Systematic Review. Artigas A, editor. Crit Care Res Pract. 2019;2019:6902420.

7. Petersen JA, Rasmussen LS, Rydahl-Hansen S. Barriers and facilitating factors related to use of early warning score among acute care nurses: a qualitative study. BMC Emerg Med. 2017;17(1):36.

8. Petersen JA, Mackel R, Antonsen K, Rasmussen LS. Serious adverse events in a hospital using early warning score. 2013; What went wrong? Resuscitation. 2014 Dec 1;85(12):1699–703.

9. Khanna AK, Hoppe P, Saugel B. Automated continuous noninvasive ward monitoring: future directions and challenges. Crit Care. 2019;23(1):194.

10. Greenhalgh T, Wherton J, Papoutsi C, Lynch J, Hughes G, A’Court C, et al. Beyond Adoption: A New Framework for Theorizing and Evaluating Nonadoption, Abandonment, and Challenges to the Scale-Up, Spread, and Sustainability of Health and Care Technologies. J Med Internet Res. 2017;19(11):e367.

11. Stafseth SK, Grønbeck S, Lien T, Randen I, Lerdal A. The experiences of nurses implementing the Modified Early Warning Score and a 24-hour on-call Mobile Intensive Care Nurse: An exploratory study. Intensive Crit Care Nurs. 2016;34:33–41.

12. Jensen JK, Skår R, Tveit B. Hospital nurses’ professional accountability while using the National Early Warning Score: A qualitative study with a hermeneutic design. J Clin Nurs. 2019 Dec 1;28(23–24):4389–99.

13. Bigham BL, Chan T, Skitch S, Fox-Robichaud A. Attitudes of emergency department physicians and nurses toward implementation of an early warning score to identify critically ill patients: qualitative explanations for failed implementation. CJEM. 2018/06/14. 2019;21(2):269–73.

14. Jensen JK, Skår R, Tveit B. Introducing the National Early Warning Score – A qualitative study of hospital nurses’ perceptions and reactions. Nurs Open. 2019 Jul 1;6(3):1067–75.

15. SmartSurvey. [cited 2022 Feb 21].

16. Braun V, Clarke V. Thematic analysis. 2012;

17. Cherry PG, Jones CP. Attitudes of nursing staff towards a Modified Early Warning System. Br J Nurs. 2015 Sep 10;24(16):812–8.

18. Jensen JK, Skår R, Tveit B. The impact of Early Warning Score and Rapid Response Systems on nurses’ competence: An integrative literature review and synthesis. J Clin Nurs (John Wiley Sons, Inc). 2018 Apr;27(7–8):e1256–74.

19. Bardi T, Gómez-Rojo M, Candela-Toha AM, de Pablo R, Martinez R, Pestaña D. Rapid response to COVID-19, escalation and de-escalation strategies to match surge capacity of Intensive Care beds to a large scale epidemic. Rev Esp Anestesiol Reanim. 2021 Jan;68(1):21–7.

20. Mitchell OJL, Neefe S, Ginestra JC, Baston CM, Frazer MJ, Gudowski S, et al. Impact of COVID-19 on inpatient clinical emergencies: A single-center experience. Resusc plus. 2021 Jun;6:100135.

21. Montagna E, Donohoe J, Zaia V, Duggan E, O’Leary P, Waddington J, et al. Transition to clinical practice during the COVID-19 pandemic: a qualitative study of young doctors’ experiences in Brazil and Ireland. BMJ Open. 2021;11(9):e053423.

22. Kamble R, Scantling-Birch Y, Samarth G, Larsson E, Maden C, Kamble R. P67 Integrated surgical teaching for juniors: surgical education during COVID-19. BJS Open. 2021;5(Supplement_1).

23. Liu S-Y, Kang XL, Wang C-H, Chu H, Jen H-J, Lai H-J, et al. Protection procedures and preventions against the spread of coronavirus disease 2019 in healthcare settings for nursing personnel: Lessons from Taiwan. Aust Crit care Off J Confed Aust Crit Care Nurses. 2021 Mar;34(2):182–90.

24. Spina S, Marrazzo F, Migliari M, Stucchi R, Sforza A, Fumagalli R. The response of Milan’s Emergency Medical System to the COVID-19 outbreak in Italy. Vol. 395, Lancet (London, England). 2020. p. e49–50.

25. Zandieh SO, Yoon-Flannery K, Kuperman GJ, Langsam DJ, Hyman D, Kaushal R. Challenges to EHR Implementation in Electronic-Versus Paper-based Office Practices. J Gen Intern Med. 2008;23(6):755–61.

26. Gui X, Chen Y, Zhou X, Reynolds TL, Zheng K, Hanauer DA. Physician champions’ perspectives and practices on electronic health records implementation: challenges and strategies. JAMIA Open. 2020 Apr 1;3(1):53–61.

