## Appendix 1 and 2 for "Implementation of a digital early warning score (NEWS2) in a cardiac specialist and general hospital settings in the COVID-19 pandemic: A qualitative study"

### (1) Survey domains, questions and responses

Domains: (i) Condition, (ii)Technology, (iii)Wider system

| | Strongly agree | Agree | Neither agree nor disagree | Disagree | Strongly disagree | Strongly agree | Agree | Neither agree nor disagree | Disagree | Strongly disagree |
|---|---|---|---|---|---|---|---|---|---|---|
| A. The information generated by NEWS2; including score and alarms; are easy to understand | 6 | 24 | 6 | 3 | 0 | 3 | 19 | 3 | 0 | 3 |
| B. The implementation of digital recording of NEWS2 in EHRs helped in recording patients' parameters and NEWS2 | 9 | 21 | 5 | 2 | 4 | 2 | 22 | 2 | 5 | 0 |
| C. The implementation of digital recording of NEWS2 in EHRs helped in improving the escalation of care when needed | 7 | 20 | 29 | 4 | 2 | 2 | 15 | 5 | 4 | 0 |
| D. The presented model of NEWS2 in the EHRs is simple and practical. (EHR Electronic health records)/ | 2 | 27 | 10 | 2 | 4 | 4 | 5 | 7 | 0 | 5 |
| E. There is a clear policy on the application of NEWS2 in patients care in your hospital | 2 | 33 | 5 | 2 | 0 | 0 | 14 | 5 | 2 | 0 |
| | often | Occasionally | Sometimes | Almost never | I don't know | often | Occasionally | Sometimes | Almost never | I don't know |
| F. How often patients have complex conditions. i.e., comorbidities, metabolically unstable, or poorly understood condition (Excluding Covid) | 25 | 4 | 10 | 0 | 2 | 22 | 0 | 2 | 0 | 0 |
| G. How often patients cared for were diagnosed with Covid-19 / | 6 | 11 | 15 | 9 | 0 | 10 | 0 | 4 | 4 | 4 |
| H. How often patients have socioeconomic factors i.e. family, income, house condition, education, affecting their prognosis | 14 | 12 | 8 | 4 | 5 | 9 | 4 | 8 | 0 | 0 |
| I. The need for help in understanding NEWS2 and who to refer to. | Senior staff/superusers in my department. | Informatics/ tech professionals | Materials or online resources | Didn't kw where to go | Didn't need help | Senior staff/superusers in my department. | Informatics/ tech professionals | Materials or online resources | Didn't kw where to go | Didn't need help |
| | 12 | 2 | 6 | 2 | 19 | 9 | 4 | 9 | 2 | 5 |
| J. Attending networking session with regard to NEWS2 and routine monitoring i.e updates, or team meetings. | Internally in the department | Organised hospital sessions | Informal discussions with colleagues | Yes, I checked the materials we have or online resources | I haven't participated or attended any updates or networking sessions | Internally in the department. | Organised hospital sessions | Informal discussions with colleagues | Yes, I checked the materials we have or online resources | I haven't participated or attended any updates or networking sessions |
| | 10 | 5 | 6 | 0 | 28 | 5 | 2 | 4 | 5 | 12 |
| K. Training offered to understand and utilise NEWS2. | Informatics staff/ tech experts/superusers | I. Manuals and resources online. | J. Orientation by ward staff or manager | K. Self-practice | | Informatics staff/ tech experts/superusers | I. Manuals and resources online. | J. Orientation by ward staff or manager | K. Self-practice | |
| | 29 | 20 | 18 | 6 | | 12 | 9 | 4 | 5 | |

**(2) Interview domains and questions**

Domains: (i)Condition (ii)Value preposition, (iii)Adopters, (iv)Organisation (v)Adaptation over time

(i)Experience of implementing NEWS2 in the clinical setting:

- Have you used NEWS2 for escalation of care while caring for patients? How common is it to receive an alarm for a patient in need for critical care?
- How do you respond to an alarm by NEWS2?

(ii)The value of NEWS2

- How valuable is NEWS2 as a tool presented to you by the developers who implemented it in EHRs?
- How Simple was NEWS2 understanding and use?
- Does it bring you the value it was supposed to? Did it improve the efficacy and safety of patients?

(iii)The adoption to utilizing NEWS2

- How did this implementation change your practice in escalating care?
- How did this change affect patients care and safety?

(iv) The organisational capacity

- How supported is early warning scores in the organisation? Do you think the hospital systems is ready for utilizing NEWS2 digitally?

(iv)Adapting over time:

- How resilient is the organisation and staff to adapt to the escalation of care using NEWS2?
- Have you faced issues around the process of escalation using NEWS2 and technology? How much scope is there to resolve that may arise over time?
